# Phenotypic Signatures of CTNNB1 Syndrome: Longitudinal Neuropsychological Outcomes and Three-Dimensional Facial Morphology

**DOI:** 10.64898/2026.01.28.26344818

**Authors:** Mercè Pallarès-Sastre, Aroa Casado, Imanol Amayra, Samuel Anguiano, Bastián Escobar-Ramírez, Mireia Andreu-Montoriol, Ona Roure-Ramis, Esther Esteban, Xavier Sevillano, Álvaro Heredia-Lidón, Rafael Pulido, Caroline E. Nunes-Xavier, Ana Rodríguez-Ramos, Sonia Bañuelos, Fabio Cavaliere, Neus Martínez-Abadías, Maitane García

**Author notes:** Correspondence concerning this article should be addressed to Mercè Pallarès-Sastre and Aroa Casado., Mercè Pallarès-Sastre: University of Deusto, Avenida de las Universidades 24, Deusto, 48007, Bilbao, Spain. (ext. 2577)., Aroa Casado: Hospital Clínic de Barcelona, Carrer de Casanova, 143, Eixample, 08038, Barcelona, Spain. Mercè Pallarès-Sastre and Aroa Casado should be considered joint first author. Mercè Pallarès-Sastre Aroa Casado Imanol Amayra Samuel Anguiano Bastián Escobar-Ramírez Mireia Andreu-Montoriol Ona Roure-Ramis Esther Esteban Xavier Sevillano Álvaro Heredia-Lidón Rafael Pulido Caroline E. Nunes-Xavier Ana Rodríguez-Ramos Sonia Bañuelos Fabio Cavaliere Neus Martínez-Abadías Maitane García.

## Abstract

CTNNB1 syndrome is a rare neurodevelopmental disorder caused by pathogenic variants in the CTNNB1 gene. Although its core clinical manifestations have been increasingly recognised, longitudinal data on cognitive, behavioural and motor trajectories remain limited, and the craniofacial phenotype has not previously been quantitatively characterised. This study provides longitudinal evidence on the cognitive, clinical and psychological profile of individuals with CTNNB1 syndrome, together with a detailed three-dimensional morphometric analysis of facial morphology.

Cognitive, clinical, psychological and neuropsychological data were collected at two time points (T0 and T1), separated by a one-year interval, using a comprehensive and standardised assessment protocol. Longitudinal analyses indicated stability across most domains, with no evidence of systematic regression. A significant improvement in gross motor functioning was observed, particularly among younger participants. Linear mixed-effects models showed that age moderated developmental change, with younger individuals exhibiting greater gains over time in gross motor skills and adaptive behaviour compared to older participants.

Three-dimensional facial morphometric analyses revealed a distinctive and statistically significant craniofacial pattern associated with CTNNB1 syndrome, independent of age and facial size. This phenotype was characterised by midfacial narrowing, reduced midface projection and mandibular retrusion. Importantly, facial shape variation was significantly associated with externalising behavioural problems and clinically relevant behavioural difficulties, suggesting a link between craniofacial morphology and behavioural severity.

This study represents the first integrated longitudinal characterisation of CTNNB1 syndrome combining neurodevelopmental follow-up with quantitative craniofacial phenotyping. The findings indicate slow but progressive improvement in specific clinical domains during childhood and adolescence, alongside relative stability in global adaptive functioning, and highlight three-dimensional facial morphology as a sensitive structural biomarker for phenotypic stratification and clinical monitoring in CTNNB1 syndrome.

**Lay summary:** This study is the first to describe how children with CTNNB1 syndrome, a rare genetic condition that leads to global developmental delays, develop over time. We also performed advanced facial analysis to look for common facial features among patients.

## Introduction

In recent years, advances in genetic testing have enabled the diagnosis of many genetic neurodevelopmental disorders (Hamdan et al., 2014; Levy et al., 2011). One such condition is the CTNNB1 syndrome, a neurodevelopmental rare disorder caused by genetic variants in the *CTNNB1* gene, encoding *β*-catenin (Kuechler et al., 2014). *β*-catenin is an essential homeostatic regulator of cell growth and development, and *CTNNB1* variants associated with the disorder, most often generated by nonsense or frameshift mutations at the *CTNNB1* gene exonic regions, are likely to display impaired biological activity (Mirosevic et al., 2022). CTNNB1 syndrome patients exhibit an heterogenous phenotypic profile; however, several main clinical manifestations are consistently present, including developmental delay or intellectual disability (ID), language and speech impairment, abnormal muscle tone, visual problems, sleep disturbances, motor delay, behavioural problems, autistic features and microcephaly, among others (Dubruc et al., 2014; Kuechler et al., 2015).

Previously, we carried out a preliminary and transversal characterisation of cognitive, adaptive functioning, motor and clinical variables of CTNNB1 syndrome patients using a novel assessment protocol that included in-person assessments and online parent-reported questionnaires (Pallarès-Sastre et al., 2025a; 2025b). Behavioural problems were more prevalent than previously reported, with around 60% exhibiting symptoms of Autism Spectrum Disorder (ASD) (Pallarès-Sastre et al., 2025a). We also detected a wide age range in which individuals reached motor and language developmental milestones, although attainment was delayed in most cases. Adaptive functioning was consistently below chronological-age expectations across all the sample, with verbal patients outperforming nonverbal ones (Pallarès-Sastre et al., 2025a). For comparative purposes, patients with ASD and Cerebral Palsy (CP) were also assessed to explore differences in cognitive and adaptive functioning. While verbal tasks performance of CTNNB1 syndrome patients was on pair with control groups, they scored significantly lower in visuospatial and logical reasoning tasks. In comparison, patients with ASD demonstrated better adaptive functioning skills, while CP patients primarily exhibited differences in gross motor abilities (Pallarès-Sastre et al., 2025a).

Age of participants was positively correlated with cognitive performance in individuals with CTNNB1 syndrome, suggesting improvement over time, with exception of language abilities, highlighting the heterogeneous language profile (Pallarès-Sastre et al., 2025b). Previous studies reported progressive but modest improvements in cognitive and motor functioning, particularly among paediatric participants (Lee et al., 2023; Pipo-Deveza et al., 2018). In fact, Sudnawa et al. (2024) provided statistical evidence suggesting continued gain of motor skills over time. However, the very few adult cases reported in the literature have shown motor regression (Tucci et al., 2014; Verhoeven et al., 2020).

Despite these advances, three critical gaps remain in the existing literature. In the first place, longitudinal data on cognitive and motor development on CTNNB1 syndrome patients remain scarce (Pallarès-Sastre, Amayra, Salgueiro et al. 2025), and there are no prospective longitudinal studies of CTNNB1 syndrome published to date. As a result, the developmental progression of symptoms in CTNNB1 it is still unclear. Natural history studies aimed at tracking the development and experiences of individuals with CTNNB1 syndrome over time are needed to further understand the progression of the disorder (Miroševič et al., 2025). Second, there is limited evidence regarding the correlation between the specific *CTNNB1* genetic variants and the phenotype presented by individuals with CTNNB1 syndrome. We recently performed an *in vitro* biochemical and functional characterisation of the *CTNNB1* variants present in these patients, and revealed an association between protein stability of the *β-*catenin variants and the patient performance in visuospatial neurocognitive tests (Nunes-Xavier et al., 2025). Third and finally, we observed that the phenotypic variability in CTNNB1 syndrome, including the facial gestalt qualitatively described in the literature (Ji et al., 2023; Yan et al., 2022; Braddock et al., 2020; Lee et al., 2025; Winczewska-Wiktor et al., 2016), appeared to be influenced by genotype, mosaicism, and population background (Pallarès-Sastre et al., 2025b).

To address these gaps, we first conducted a longitudinal follow-up of the trajectory of global cognitive abilities, adaptive skills and clinical variables on a group of CTNNB1 syndrome patients, to start exploring the natural history of this ultrarare disorder. Second, we quantitatively assessed whether there is a facial dysmorphology pattern associated with CTNNB1 syndrome. Several studies have reported that individuals with CTNNB1 syndrome present typical facial features as a broad and flat nasal bridge, bulbous nasal tip, and downturned mouth corners, among other features (Ji et al., 2023; Yan et al., 2022; Braddock et al., 2020; Lee et al., 2025; Winczewska-Wiktor et al., 2016). However, no previous study has directly quantified the facial pattern of CTNNB1 syndrome.

Considering that up to 40% of rare genetic disorders show characteristic facial patterns due to the sensitivity of facial morphogenesis to genetic alterations (Martínez-Abadías et al., 2011; Hallgrímsson et al., 2020), and the limitations of automated facial phenotyping tools in the context of ultrarare diseases (Gurovich et al., 2019; Echeverry-Quiceno et al., 2023; Alvi et al., 2022; Wadden, 2022; Hsieh et al., 2022; Hustinx et al., 2023), the implementation of standardised and reproducible methodologies becomes essential. To address this aim, we employed a high-resolution multicamera photogrammetry protocol and a morphometric pipeline based on validated landmarks (Heredia-Lidón et al., 2025; Gunz et al., 2005; Gunz & Mitteroecker, 2013). We then used geometric morphometric methods to comparing the three-dimensional facial morphology of individuals with CTNNB1 syndrome with sex and age matched controls. Finally, we tested whether these precise facial shape measurements were associated with the clinical, psychological and neuropsychological variables assessed in the longitudinal cognitive analysis in order to identify which clinical dimensions exhibit a detectable morphological signature.

Therefore, this study not only provides a longitudinal cognitive and psychological characterisation of CTNNB1 syndrome patients, but provides a detailed description of facial morphology based on a relatively large paediatric cohort of individuals with CTNNB1 syndrome, and evaluates its potential to function as a complementary phenotypic indicator of the clinical domains characterised longitudinally.

## Methods

### Participants and procedure

The patient sample consisted of 25 Spanish children diagnosed with CTNNB1 syndrome, including 16 females and 9 males from 3 to 18 years old, recruited through the CTNNB1 Spanish Association (Asociación CTNNB1 España). For the clinical and cognitive characterisation, parents of children with CTNNB1 syndrome were also recruited. For assessing facial morphology in these children diagnosed with CTNNB1 syndrome, we recruited a comparative sample of sex and age controls at data collection sessions organized at Barcelona, Burgos, Bilbao and Terrassa (Table 1).

The inclusion criteria followed for patients were: a) to have a genetic diagnosis of CTNNB1 syndrome, b) to have been assessed with the cognitive and psychological measures in T0, c) to have Spanish as one of the primary languages, and d) to have the Spanish nationality. The exclusion criteria for patients was to have other diagnoses not secondary to CTNNB1 syndrome. The associated patient genetic data can be found in Pallarès-Sastre et al. (2025a). Clinical history of craniofacial trauma and/or surgery were exclusion criteria for all participants.

All study procedures were in line with the Declaration of Helsinki, complying with all relevant ethical regulations and the protocol was approved by the Ethics Committee of the University of Deusto (ETK-24/23-24) and the Bioethics Committee of the Universitat de Barcelona (IRB03099-CER032514). All individuals, including parents or legal guardians in case of minors, provided informed consent prior participation.

### Clinical, cognitive, adaptive, and behavioural longitudinal characterisation

This study followed a longitudinal approach, with data collected at two time points separated by one year; the baseline data (T0) were conducted from November 2023 to June 2024, followed by the second assessments one year later (T1). Participants underwent in-person cognitive assessments, which were carried out by the same neuropsychologists who travelled across the country during family meeting. These sessions were dived in two parts of half an hour each to guarantee the patient’s attention. Parent-reported questionnaires were assessed through the software *Qualtrics* and online interviews with a neuropsychologist for hetero-applied measures. Finally, parents were asked to request their child’s physiotherapist to assess the GMFM88 with prior consent of the professional. T0 data is already published in Pallarès-Sastre et al., (2025a) and Pallarès-Sastre et al., (2025b).

## Instruments

### Sleep Problems

#### Sleep disturbances Scale for children

(Pagerols et al., 2023). The aim of this scale is to provide a comprehensive measure of sleep disturbances for clinic and research purposes. It includes 26 Likert-type items and it is divided into five subdomains: disorders of initiating and maintaining sleep, sleep breathing disorders, disorders of arousal, sleep-wake transition disorders, and disorders of excessive somnolence and sleep hyperthyroidism. A total score of 39 or higher indicates the presence of a global sleep disorder and it is answered by the parents.

### Ability to eat

#### Eating and Drinking Ability Classification System

(EDACS) (Sellers et al., 2013). This instrument classifies individuals on the basis of how efficiently individuals eat and drink in their daily lives using a five-level scale; from drinking and eating safely to unable to do so. The parents are asked to choose the description that bests suits the ability of their child.

### Communication

#### Viking Speech Scale

(VSS) (Lindsay et al., 2013). The purpose of this scale is to evaluate children’s speech production based on the parent’s knowledge, specifically focusing on how understandable their speech is. It includes four level options, ranging from unaffected speech to not understandable speech.

#### Communication Function Classification System

(CFCS) (Keith & Arellano, 2012). This test provides five levels to describe everyday communication performance by the parent’s perspective. It assesses the child’s ability to engage in conversations where the roles of sender and receiver of information are being exchanged.

#### Peabody Picture Vocabulary Test, Third Edition

(PPVT-3) (Dunn et al. 2010). The aim of this instrument is to assess receptive language and it is used as a screening tool for verbal skills. The examinee has to choose the picture that best matches the word spoken by the examiner, out of four simple black-and-white options. The items are organized into 17 sets of 12, arranged in order of increasing difficulty.

#### The Boston Naming Test

(BNT) (Kaplan et al. 2005). This instrument assesses lexical ability using black and white illustrations of increasing difficulty. If the examinee is unable to name the image, the examiner provides phonemic or phonetic clues and, if necessary, offers multiple-choice options as a final prompt.

### Motor Functioning

#### Manual Ability Classification System for Children with Cerebral Palsy

(MACS and mini-MACS) (Eliasson et al., 2006). The purpose of this test is to ask the parents/caregivers which of the five levels best describe the ability of their child when using their hands for handlining objects. The original version, MACS, is designed for children aged 4 to 18 years, while the mini-MACS is and adapted version for children ages 1 to 4 years.

#### Gross Motor Function Measure 88

(GMFM88) (Ferre-Fernández et al., 2020). The GMFM88 is widely used to evaluate gross motor functioning in children with cerebral palsy and it is divided into five dimensions: lying and rolling, sitting, crawling and kneeling, standing and walking, running and jumping. Scores range from zero to three for each item conforming the dimensions, from which a percentage is calculated. Also, the tests offer an overall percentage of the gross motor functioning.

### Neurodevelopment

#### Vineland Adaptative Behavior Scales-3

(Sparrow et al., 2016). The Vineland-3 is a hetero-applied measure with the parents/caregivers that measures adaptive functioning of individuals with intellectual and development disabilities. The instrument evaluates five main domains: Communication (COM), Daily Living Skills (DLS), Socialisation (SOC) motor skills and maladaptive behaviour. Each domain includes several subdomains and, for each subdomain, a raw score is obtained and then converted into a V-scale score, which is calculated based on the examinee’s age. Scores less than 70 are considered impaired (70-79 borderline; 80-89 below average; 90+ average). The Internalising and Externalising subdomains from the maladaptive scale can also be clinically interpreted using categories that range from symptoms within normal limits, moderately elevated and clinically significant.

### Autism

#### Form B of the Social Communication Questionnaire

(SCQ) (Pereña & Santamaría, 2005). This instrument is a parent-reported questionnaire to evaluate three core aspects of ASD: social relating, communication and range of interest. The purpose is to determine whether the child exhibits symptomatology consistent with ASD. Consists of 40 yes-or-no items and a score higher than 15 indicates possible compatibility with ASD symptoms.

#### Childhood Autism Rating Scale

(CARS) (García-Villamisar, 1992). The aim of the CARS is to detect early onset of symptoms related to ASD. It is structured as a parent-completed questionnaire of 15 items, each rated from 1 to 5, resulting in a total score range of 15 to 60. Depending on the scores, participants can be classified into three groups; minimal-to-no symptoms of ASD, mild-to-moderate symptoms of ASD and severe symptoms of ASD.

### Cognitive measures

#### Wechsler Nonverbal Scale of Ability

(WNV) (Wechsler & Naglieri, 2011). The WNV is a non-verbal neuropsychological battery that assess cognitive development of children with language impairments, autism or hearing difficulties. For this study, only three of the six subtests were administered. First, the Matrices subtest assess perceptual reasoning through simultaneous processes. Additionally, the Object Assembly subtest measures perceptual organisation and reasoning. Finally, the Recognition subtest evaluates immediate memory through visual stimulus.

### Comprehension of instructions

(CI) from the NEPSY-II (Korkman et al. 2017). This subtest evaluates the ability to receive, process and execute oral instructions of increasing syntactic complexity. After hearing the oral instruction, the examinee has to point the correct stimuli.

### Statistical analyses

Statistical analyses for assessing clinical, cognitive, adaptive, and behavioural longitudinal characterisation were conducted using SPSS version 28.0. Given the limited sample size, non-parametric tests were applied. Descriptive statistics were computed for all T0 and T1 variables, and scores were transformed into *Z*-scores. Longitudinal differences between both time points were examined using the Wilcoxon signed-rank test. To assess whether age predicted within-subject variation in cognitive and psychological outcomes across the one-year interval, linear regression analyses and linear mixed-effects models (LME4 package, R) were conducted.

### Facial Morphometry

#### Facial Data Acquisition and 3D Model Reconstruction

Facial images were acquired using a stereophotogrammetry system consisting of ten Sony Alpha A58 DSLR cameras mounted on five tripods arranged along a semicircle, positioned one meter from the participant’s face with an overlap angle below 30°. This multi-camera configuration enabled the simultaneous capture of multiple perspectives of the face, ensuring optimal coverage for high-resolution three-dimensional reconstruction. To minimize artefacts caused by dynamic facial expressions, all photographs were taken with participants in an upright seated position, looking straight ahead with open eyes and closed lips. In younger participants with CTNNB1 syndrome disorder, who often exhibited difficulty maintaining a stable posture or showed lingual protrusion, multiple photographs were taken until a neutral facial expression was obtained. The final selection of the ten optimal images per participant was performed using *PhotoSelector* (proprietary software). High-resolution 3D facial meshes were generated using Photo3DFace, a proprietary software based on Agisoft Metashape 2.2.0 (Agisoft LLC, 2024). This pipeline yielded calibrated and scaled facial models following the procedures validated by Heredia-Lidón et al. (2025).

A total of 21 anatomical landmarks (Figure 1; Supplementary Table 1) were automatically recorded on each 3D model using the facial detection module of Photo3DFace. This system employs a multi-view consensus convolutional neural network (MV-CNN) architecture (Paulsen et al., 2019), an advanced deep learning approach for automatic anatomical landmarking. The algorithm generates multiple 2D projections of the facial mesh from different viewpoints, uses an hourglass CNN model to predict heatmaps for potential landmark locations, and determines the final 3D coordinates through least-squares ray-projection with a random sample consensus procedure (Paulsen et al., 2019). The model was trained on 125 manually landmarked facial meshes. All automatic landmark placements were reviewed and, when necessary, corrected by a single trained observer using 3D Slicer (Fedorov et al., 2012).

**Figure 1.**
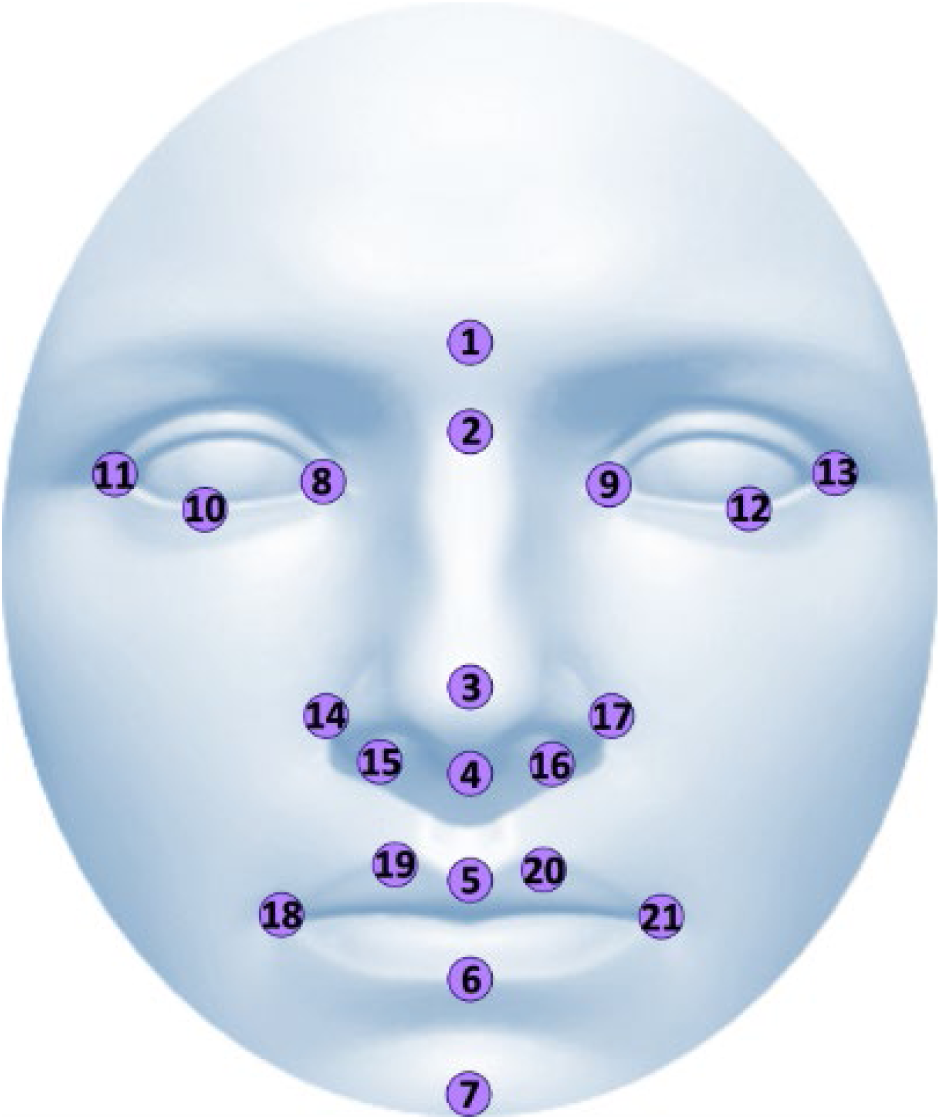
*Note*. 3D facial reconstruction showing the 21 anatomical landmarks (numbered symbols). Landmark definitions are provided in Supplementary material 1.

### Multivariate Morphometric and Statistical Analyses

To analyse facial shape variation, we applied Geometric morphometrics (GM), a robust set of statistical tools designed for measuring and comparing 3D shapes with high precision and efficiency (Bookstein, 1992; Dryden & Mardia, 2016; Hallgrímsson et al., 2015). Facial landmark configurations were aligned to a common morphospace using Generalised Procrustes Analysis (GPA), removing non-shape variance due to translation, rotation, and scale. Facial size was quantified as centroid size, defined as the square root of summed squared distances from landmarks to the centroid (Dryden & Mardia, 2016). The resulting Procrustes coordinates served as the basis for all morphometric analyses.

First, to assess whether CTNNB1 syndrome diagnosis and other variables such as sex, age and size were significantly associated to facial shape variation, we performed a permutation-based ANOVA test using RRPP with 10,000 permutations (Adams et al, 2013; Larson et al, 2018), as implemented with the ProcD.lm function from the Geomorph R package (version 4.0.8).

Then, we performed a multivariate linear regression of facial shape on facial size to quantify the allometric component of facial variation, and used the regression residuals to further explore facial non-allometric differences between CTNNB1 syndrome patients and matched controls using Principal Component Analysis (PCA). Morphological variation was visualized in the morphospace defined by the first two principal components (PCs). Group differentiation was assessed using Procrustes distances between the mean shape of patient and control groups, calculated as the square root of the summed squared differences between homologous landmarks (61). Statistical significance was determined through 1,000-permutation tests.

To visualize facial shape differences between CTNNB1 syndrome patients and controls, we conducted a Canonical Variate Analysis (CVA), a statistical discriminant method that maximizes the separation among predefined groups (Campbell et al, 1981; Klingenberg et al, 2005). Facial morphings associated with the negative and positive extremes of the first two CVs were created with IDAV Landmark Editor version and MorphoJ version 1.07a b (Klingenberg et al, 2011), respectively. Moreover, we created heatmaps using Amira 5.2.1 (v6.3, Visualization Sciences Group, FEI) to identify the facial feature where differences between diagnostic groups were mainly located.

Finally, we integrated the phenotypic analysis between facial, clinical and neuropsychological biomarkers in CTNNB1 syndrome patients by performing multivariate linear regressions between facial shape variation and the variables assessing all clinical, psychological and neuropsychological variables at T1.

## Results

### Clinical longitudinal information

Table 2 shows the results of the longitudinal analysis for clinical, psychological and neuropsychological measures. Most of the results were not significant, in exception of the total score of GMFM-88, represented in Figure 2a, with a significant improvement of motor functioning in the younger population of the sample.

**Figure 2.**
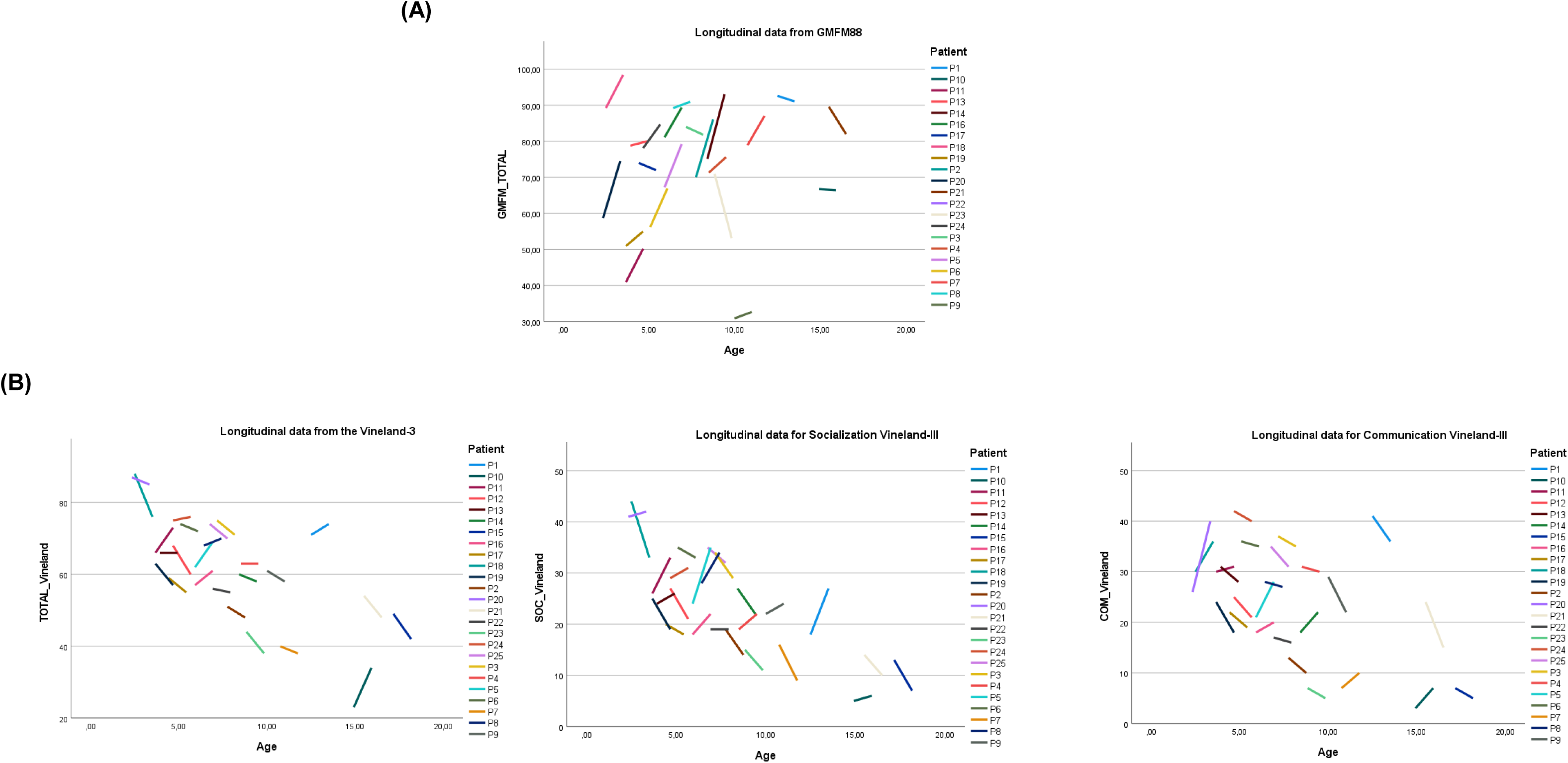
*Note*. Longitudinal graphs of the outcome measure from T0 and T1 in the CTNNB1 syndrome sample.

The graphics in Figure 2b show a visual pattern suggesting that older patients, specifically adolescents, obtain lower scores in overall adaptive functioning, as well as in the socialisation and communication subdomains. In contrast, within-subject test analyses have showed no significant difference over time (T0 vs T1) (Table 2). Specifically, Figure 3 shows a comparison of the normative results of the domains of Vineland-III between T0 and T1, which further support the stability of the results from a clinical perspective.

**Figure 3.**
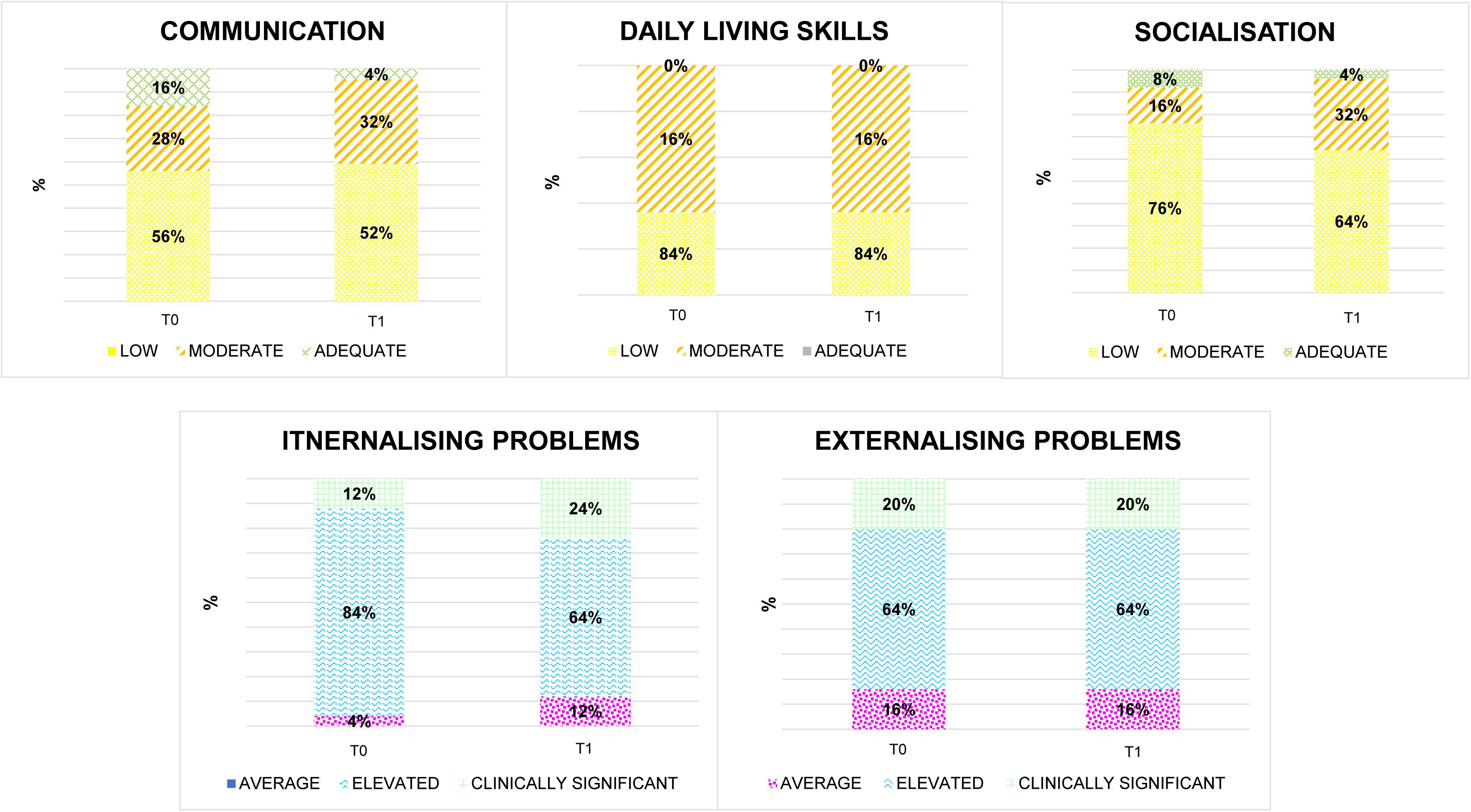
*Note*. Clinical interpretation of the normative scores for the Vineland-3 domains at T0 and T1.

A linear mixed-effects model was conducted to examine whether age moderated change in certain variables. The model included T0 and T1 and the ages of participants, in order to assess the interaction between the variables. Results were interpreted based on the direction and magnitude of effects rather than strict statistical thresholds. The first one, was the sum of the raw scores of all subdomains from the Vineland-3, to analyse whether the amount of change depended on age. The main effect of age was positive, and the negative Time × Age interaction (*B*= –4.45, *t*= –2.0) indicated that younger participants showed greater gains over time than older ones. The second analysis included the GMFM88 outcomes from T0 and T1. The main effect of age was not significant but the Time × Age interaction was negative and statistically significant (*B*= -0.94, *t*= -2.03), so that younger participants proved greater improvements compared to older ones in gross motor functioning over time (Figure 4).

**Figure 4.**
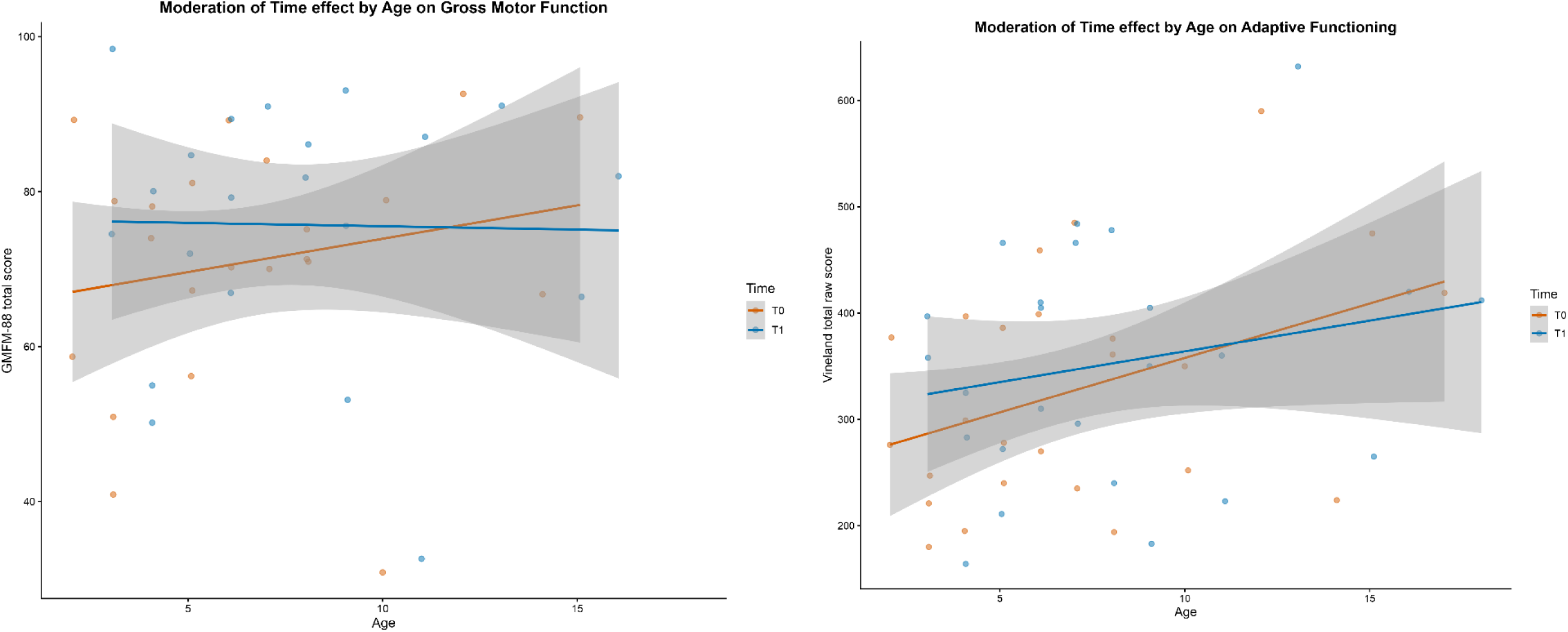
*Note*. Moderation of Time by Age on Vineland adaptive functioning and gross motor functioning. Scatter points represent individual participants at T0 and T1 assessments. Solid lines depict fitted linear trends, with shaded bands indicating 95% confidence intervals.

During this year, we collected data regarding the therapies and frequencies followed by the patients from T0 to T1 (Figure 5). The most attended therapy was physiotherapy (*N*= 23), followed by speech therapy (*N*=22) and occupational therapy (*N*=14). Table 3 summarizes the main goal of each therapy in the recruited samples based on the parent’s perspectives.

**Figure 5.**
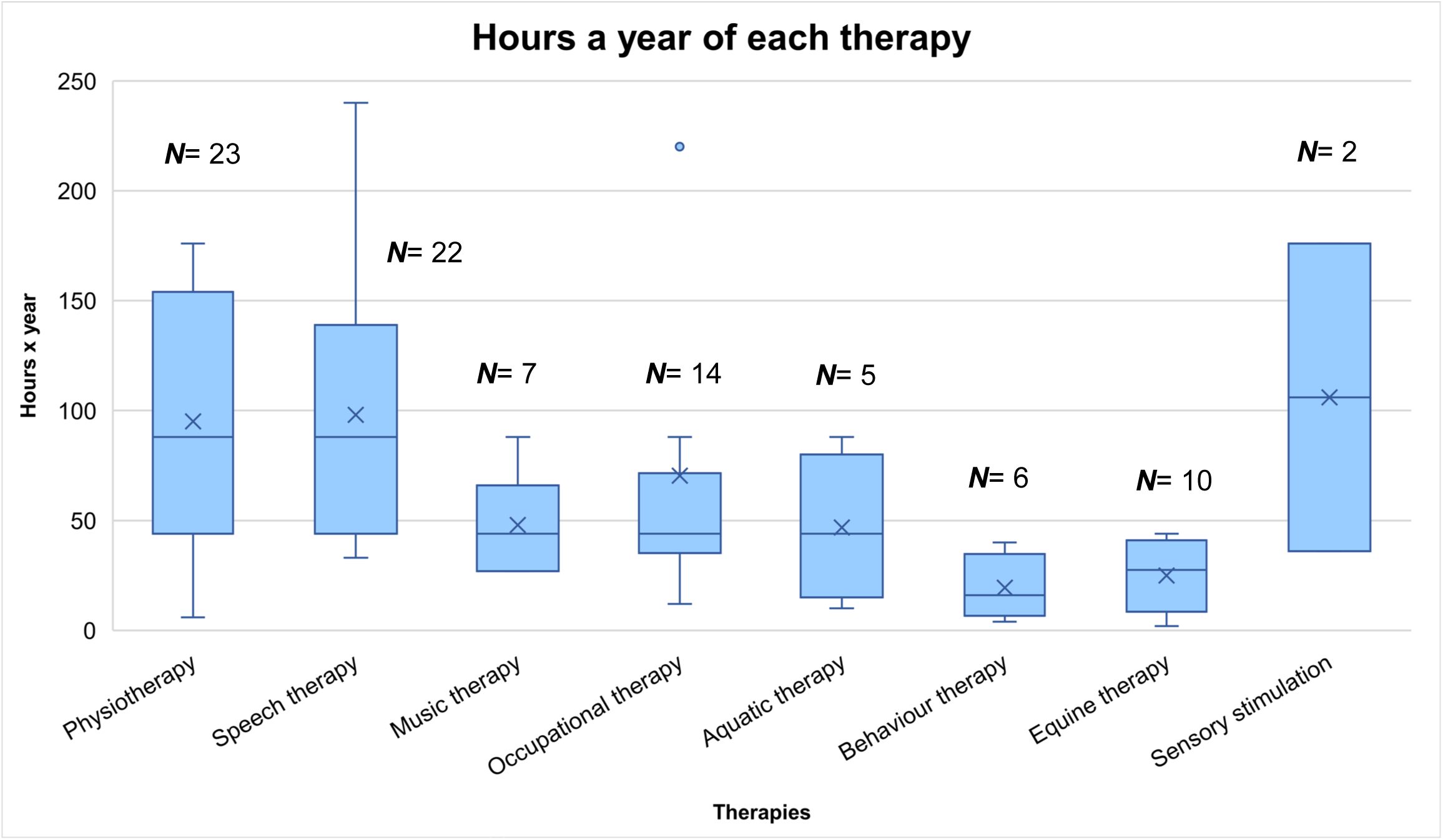
*Note*. Number of patients that have attended each therapy and hours a year accumulated in each therapy by all participants.

### Morphological Analyses

Regarding the face, the ANOVA Procrustes analysis showed that CTNNB1 syndrome diagnosis explained a significant percentage of facial shape variation (*p*= 0.032). This result was obtained, after accounting for the variation that is due to age and size differences between the individuals in the sample (Table 4), which significantly explained 9.9 and 14.1% of facial shape variation (*p*= 0.0001), respectively. As expected for a paediatric cohort of young children, sex was not a significant factor for facial shape variation (Table 4).

The Procrustes distance between patient and control groups further underscored that CTNNB1 syndrome is associated with a significant and unique facial pattern (Procrustes distance= 0.0398; *p*= 0.002). Again, this difference was maintained after removing the allometric effect, and the facial shape differences associated with growth (Procrustes distance= 0.0358; *p*= 0.0007). CTNNB1 syndrome patients showed a significantly different facial morphology, including narrower faces, a slight midface projection and chin retrusion (Figure 6).

**Figure 6.**
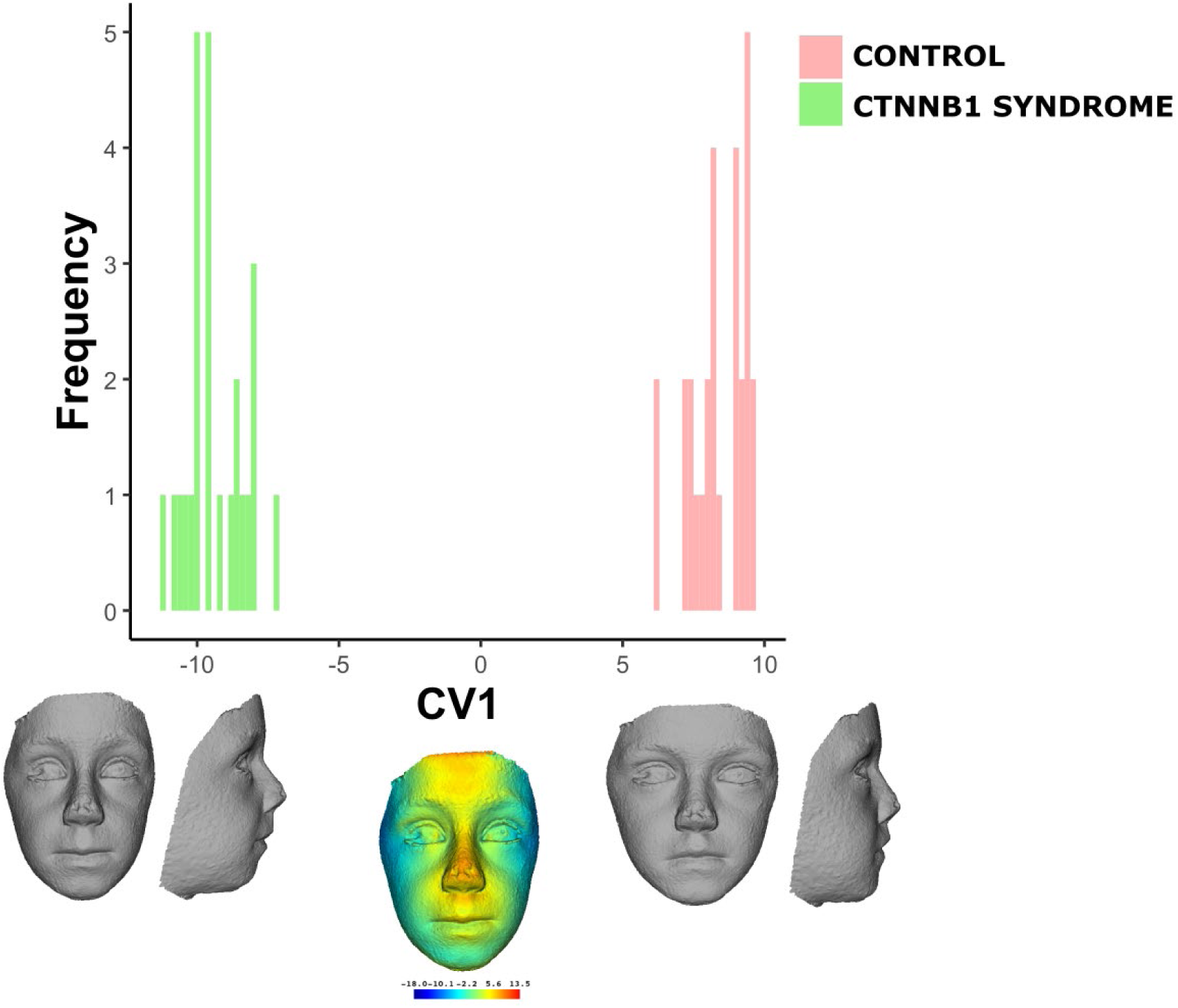
*Note*. CVA scatterplot based on facial shape variation in individuals with CTNNB1 syndrome. Morphings represent facial morphology at the positive (control) and negative extremes of CV (CTNNB1 syndrome). Heatmap shows facial regions that present the highest variation along CV1. The faces in the figure are morphings which do not belong to any real individual and are created from 3D landmark coordinates.

The multivariate linear regressions assessing the integration of the facial phenotype with clinical and neuropsychological and psychological variables only reported a significant association between facial shape and two variables: externalising problems (*p*= 0.002) and critical problem behaviours (*p*= 0.01) (Figure 7), which were moderately associated among them, as revealed by a Pearson’s correlation test (*r*= 0.44; *p*= 0.037). Individuals with highest scorings of externalising and critical problem behaviours showed more pronounced CTNNB1 syndrome facial features (Figure 7).

**Figure 7.**
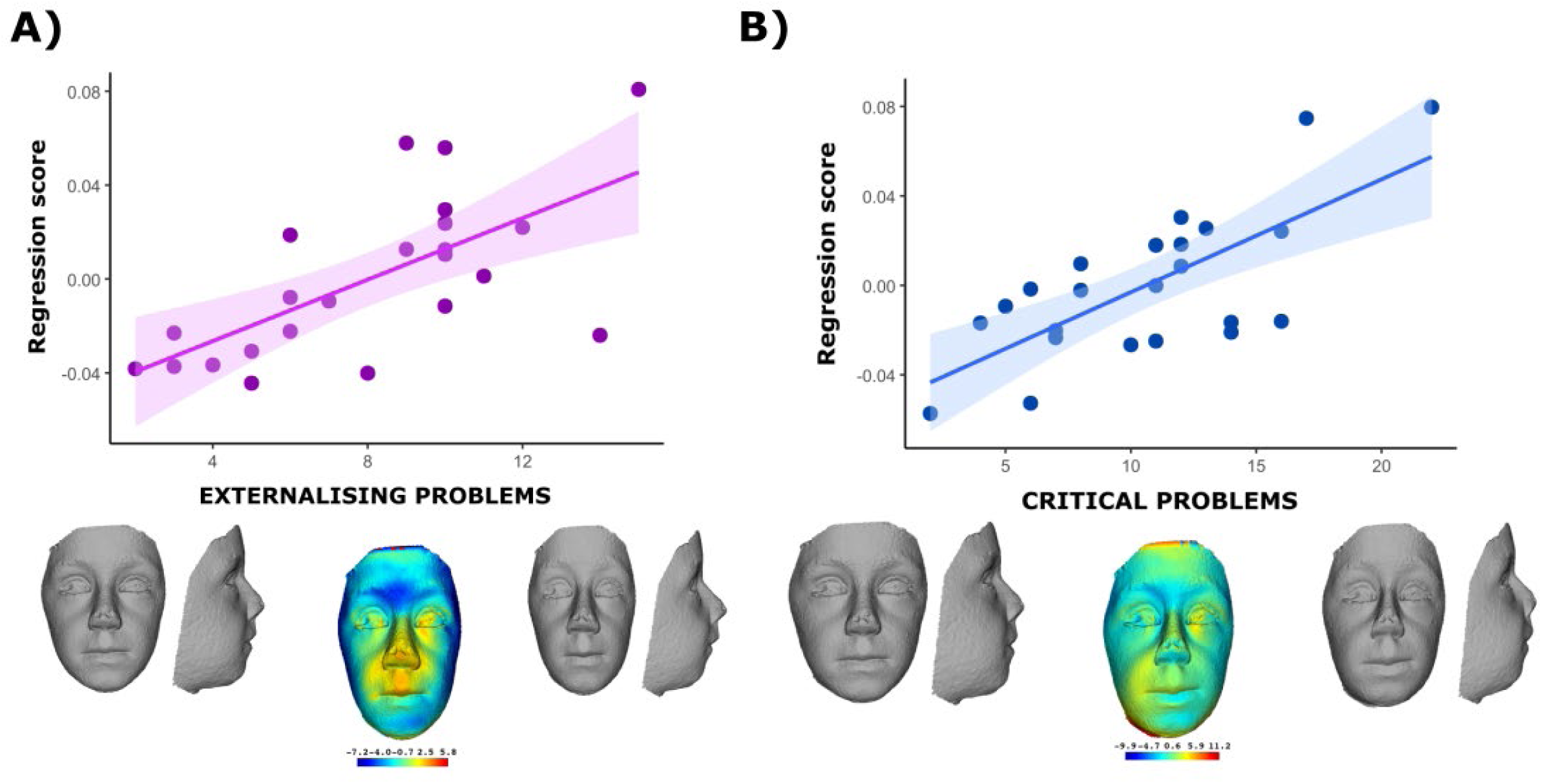
*Note*. Multivariate linear regressions of externalising problems (A) and critical problem behaviours (B) on the facial shape variation of individuals with CTNNB1 syndrome. Morphings show frontal and lateral facial shape changes of the severity extremes. Heatmaps indicate regions of greatest variation. The faces in the figure are morphings which do not belong to any real individual and are created from 3D landmark coordinates.

Besides externalising and critical problem behaviours, no other significant associations were detected with the rest of clinical, psychological and neuropsychological variables. Only the association with the Peabody-3 test assessing receptive language was marginally significant (*p*= 0.08).

## Discussion

To our knowledge, this study provides the most comprehensive longitudinal characterisation to date of neurodevelopmental, behavioural, motor and facial variation in individuals with CTNNB1 syndrome. By integrating in-person cognitive assessments, parent-reported questionnaires, motor evaluations and quantitative three-dimensional facial morphometry, this work offers a multimodal perspective on short-term developmental trajectories. Twenty-five participants were assessed at baseline and re-evaluated one year later, addressing a critical gap in the literature, as longitudinal data in CTNNB1 syndrome remain scarce. Moreover, no previous study has examined facial morphology in relation to longitudinal neurodevelopmental or behavioural features, placing the present work in a unique position to characterise both stability and variability across phenotypic domains affected by *CTNNB1* haploinsufficiency (Dubruc et al., 2014; Ho et al., 2022).

### Stability and age-related effects on adaptive functioning

Although no statistically significant changes were observed in the main Vineland-3 adaptive behaviour domains, the apparent downward trend in normative scores is most plausibly explained by the well-described age-related effect inherent to norm-referenced adaptive measures. In individuals with persistent developmental immaturity, adaptive skills may continue to develop in absolute terms while failing to keep pace with age-matched norms, resulting in progressively lower standard scores over time. This phenomenon has been consistently documented in neurodevelopmental conditions associated with intellectual disability and autistic traits, where adaptive functioning tends to plateau relative to normative expectations rather than showing true developmental regression (Schatz, & Hamdan-Allen, 1995; Bordes Edgar et al., 2024). From a clinical standpoint, distinguishing stagnation relative to age norms from genuine loss of previously acquired skills is essential for accurate interpretation of longitudinal trajectories and for appropriate counselling of families.

Despite the absence of change in Vineland-3 composite domains, improvements in expressive and written communication, together with increased speech intelligibility, suggest that individuals with CTNNB1 syndrome may continue to acquire specific cognitive and communicative skills during late childhood and adolescence, even in the context of limited global adaptive growth. This pattern is consistent with recent clinical descriptions of CTNNB1 neurodevelopmental disorder reporting heterogeneous developmental gains across language and reasoning domains (Lee et al., 2023; Pallarès-Sastre et al., 2025a) and highlights the importance of domain-specific longitudinal assessment beyond global adaptive indices.

### Longitudinal variability and developmental timing

The literature addressing developmental trajectories in CTNNB1 syndrome remains limited and heterogeneous. Parental reports from transversal studies have described apparent regression in early language or motor abilities in a subset of individuals; however, such observations have not been consistently corroborated by objective longitudinal assessments (Žakelj et al., 2025). In contrast, detailed clinical characterisations have documented continued improvement in gross motor functioning and variable cognitive progression over time (Sudnawa et al., 2024). Additional evidence indicates that the age at which early developmental milestones are achieved is strongly associated with later domain-specific performance, underscoring the importance of early developmental timing as a determinant of long-term functional outcomes (Pallarès-Sastre et al., 2025b).

Evidence regarding therapeutic interventions in CTNNB1 syndrome remains scarce. To date, no controlled clinical trials have evaluated pharmacological or rehabilitative treatments. Isolated case reports, such as the use of L-dopa to address persistent hypertonia, illustrate potential avenues for symptomatic management but require systematic evaluation within longitudinal or interventional frameworks (Algethami et al., 2025).

In adulthood, available data are extremely limited and restricted to isolated but clinically informative case reports describing progressive motor decline and increasing spasticity leading to loss of ambulation in mid-to-late adulthood (Tucci et al., 2014; Verhoeven et al., 2020). Comparable late-emerging deterioration has been reported in other neurodevelopmental disorders, including Rett syndrome and Down syndrome (Foley et al., 2011; Bisgaard et al., 2021), highlighting the need for lifespan-oriented follow-up in CTNNB1 syndrome.

### A robust and clinically sensitive craniofacial phenotype

One of the major contributions of the present study lies in the integration of three-dimensional facial morphology as a quantitative marker of clinical phenotype. Our findings confirm a unique and recognisable craniofacial pattern in CTNNB1 syndrome, characterised by a narrow or flattened nasal bridge, reduced infraorbital convexity, thin upper lip and mandibular retrusion. These features are consistent with prior qualitative descriptions in clinically characterised cohorts and reinforce the existence of a coherent facial phenotype associated with CTNNB1 haploinsufficiency (Kuechler et al., 2015; Yan et al., 2022; Ji et al., 2023; Ho et al., 2022).

Comparisons with neurotypical controls revealed systematic reductions in midfacial projection, perioral volume and mandibular prominence, supporting the view that facial morphology constitutes a sensitive structural marker capturing both the basal phenotype of CTNNB1 syndrome and inter-individual neurodevelopmental variability. The application of geometric morphometric techniques enables objective, observer-independent quantification of these traits and provides a robust framework for phenotypic comparison (Adams & Otárola-Castillo, 2013; Bardua et al., 2019).

### Biological interpretation of the association between facial morphology and behavioural severity

The association between facial morphology and behavioural features, particularly externalising problems and clinically significant behavioural difficulties, can be interpreted within a developmental and neurogenetic framework centred on the pleiotropic role of *CTNNB1*. The *CTNNB1* gene encodes *β*-catenin, a multifunctional protein acting as a central effector of canonical Wnt/*β*-catenin signalling and as a key component of cadherin-mediated cell adhesion. *β*-catenin plays critical roles in neural crest induction, craniofacial morphogenesis and forebrain development, providing a shared biological substrate for facial and neurodevelopmental phenotypes (Chenn, 2008; Ji et al., 2019; Ho et al., 2022).

Canonical Wnt signalling is essential at multiple stages of neural crest development and regulates the induction and specification of neural crest derivatives that contribute extensively to craniofacial structures (Sutton et al., 2021; Elkouby & Frank, 2010). Disruption of these early morphogenetic programs offers a plausible explanation for the craniofacial phenotype observed in CTNNB1 syndrome. Importantly, Wnt/*β*-catenin signalling also plays a fundamental role in central nervous system development, including forebrain patterning and neuronal specification, providing a mechanistic basis for the co-occurrence of craniofacial and neurodevelopmental alterations (Brault et al., 2001; Choe et al., 2014; Marchini et al., 2021).

Beyond early morphogenesis, *β*-catenin contributes to synaptic organisation, stability and plasticity, with downstream effects on the maturation of neural circuits involved in behavioural regulation (Kwiatkowski & Nigh, 2007; Maguschak & Ressler, 2012; Tang, 2014). Experimental disruption of *CTNNB1* in animal models preferentially affects inhibitory circuitry and results in behavioural phenotypes characterised by impulsivity and dysregulation, closely aligning with externalising behavioural profiles observed in neurodevelopmental disorders (Dong et al., 2016). Together, these findings provide a biologically grounded link between CTNNB1 haploinsufficiency, altered circuit-level development and behavioural regulation.

From a functional perspective, externalising behaviours depend on the integrity of fronto-limbic and fronto-striatal networks that undergo prolonged postnatal maturation and are particularly sensitive to early synaptic perturbations (Ameis et al., 2014). In contrast, global cognitive and adaptive measures represent composite constructs influenced by multiple neurodevelopmental, environmental and interventional factors, potentially attenuating direct associations with early morphogenetic markers.

### Comparative phenotype, convergence and specificity

The craniofacial features identified in CTNNB1 syndrome partially overlap with those described in other neurodevelopmental conditions characterised by facial dysmorphology, including fetal valproate spectrum disorder and specific chromosomal disorders, with shared features such as reduced nasal projection and a thin upper lip (DiLiberti et al., 1984; Winter et al., 1987; Clayton-Smith et al., 2019). These convergences suggest shared disruption of developmental pathways affecting neural crest derivatives and craniofacial patterning (Marcucio et al., 2015). At the same time, the specific combination of midfacial narrowing, infraorbital flattening and mandibular retrusion observed in CTNNB1 syndrome refines its phenotypic specificity and reinforces the potential of quantitative facial metrics as non-invasive biomarkers with clinical relevance.

Inter-individual variability in phenotypic expression is a well-documented feature of CTNNB1 syndrome. Wide genotypic and phenotypic variability has been reported even among individuals with similar pathogenic variants, suggesting that mutation-specific effects and additional modifying factors contribute to clinical severity and phenotypic diversity (Miroševič et al., 2022). Integrating genotypic, morphometric and longitudinal developmental data will therefore be essential for elucidating genotype–phenotype relationships and advancing precision phenotyping in CTNNB1 syndrome.

### Limitations and future directions

Several limitations inherent to the study design should be acknowledged. First, although the present cohort represents one of the largest longitudinally characterised samples reported to date in CTNNB1 syndrome, the sample size remains constrained by the rarity of the condition. This limitation reduces statistical power, particularly for detecting subtle associations across phenotypic domains, and limits the generalisability of the findings. As is common in research on rare neurodevelopmental disorders, replication in independent cohorts will be essential to confirm the robustness of the observed patterns.

Second, the one-year follow-up interval, while sufficient to capture short-term developmental changes, may be insufficient to detect slower, non-linear or late-emerging trajectories, particularly in domains such as adaptive functioning, behavioural regulation or motor progression. Longer longitudinal follow-up across multiple developmental stages will be necessary to characterise the natural history of CTNNB1 syndrome more comprehensively and to distinguish transient developmental fluctuations from sustained trajectories.

Third, although a broad battery of cognitive, behavioural and motor assessments was employed, some instruments are norm-referenced and may not be optimally calibrated for individuals with severe neurodevelopmental impairment. As discussed, age-related effects inherent to standardised adaptive measures can complicate longitudinal interpretation. Future studies would benefit from incorporating complementary assessment strategies, including criterion-referenced measures and ecologically valid functional outcomes, to better capture clinically meaningful change.

In addition, the present cohort was predominantly paediatric, limiting inferences regarding adult outcomes and lifespan trajectories. Available evidence in adulthood remains restricted to isolated case reports, and systematic longitudinal data across adolescence and adulthood are lacking. Expanding recruitment to include older individuals will be crucial for understanding long-term motor, behavioural and adaptive outcomes, as well as potential late-emerging complications associated with CTNNB1 syndrome.

Regarding intervention, although information on ongoing therapies was collected, the study was not powered to model the effects of specific therapeutic approaches on developmental trajectories. Heterogeneity in the type, intensity and duration of interventions further limits causal inference. Future longitudinal studies incorporating standardised intervention data and treatment-response measures will be essential to evaluate therapeutic impact and to inform evidence-based clinical management.

Finally, while this study represents the first quantitative analysis of facial morphology in CTNNB1 syndrome and its relationship with behavioural features, the cross-sectional nature of the morphometric–behavioural associations precludes causal interpretation. Longitudinal facial morphometric data, ideally combined with neuroimaging and neurophysiological measures, would allow for a more direct examination of how early morphogenetic variation relates to subsequent neural circuit development and behavioural outcomes.

Future research should therefore prioritise multicentre, longitudinal designs integrating genetic, morphometric, neurodevelopmental and neurobiological data across the lifespan. The combination of three-dimensional facial morphometry with neuroimaging, molecular genetic profiling and repeated behavioural assessments holds particular promise for advancing precision phenotyping in CTNNB1 syndrome and for informing pathway-oriented therapeutic strategies.

### Conclusions

This study provides the most comprehensive longitudinal characterisation of CTNNB1 syndrome to date, integrating cognitive, adaptive, motor and three-dimensional craniofacial assessments within one of the largest cohorts described for this rare neurodevelopmental condition. The findings indicate that, while global adaptive functioning remains relatively stable over a one-year period, specific domains—most notably expressive communication and speech intelligibility—continue to improve throughout adolescence. The modest downward trends observed in standardised adaptive scores are most plausibly explained by age-related norm-referencing effects rather than true developmental regression, underscoring the need for cautious interpretation of standardised adaptive measures in this population.

Three-dimensional facial morphology emerged as a sensitive structural biomarker capable of objectively capturing the core craniofacial phenotype associated with CTNNB1 haploinsufficiency. The observed associations between facial morphology and specific behavioural dimensions support the notion that the developmental pathways disrupted by *CTNNB1* haploinsufficiency exert coordinated effects on craniofacial morphogenesis and neurobehavioural functioning.

Although longitudinal evidence in CTNNB1 syndrome remains extremely limited, the present findings are consistent with previous reports describing sustained motor gains and the absence of systematic neurocognitive regression during childhood and adolescence. However, the scarcity of data in adulthood represents a critical gap, particularly in light of emerging case reports suggesting the possibility of late-onset motor decline.

Despite limitations related to sample size and clinical heterogeneity, this study establishes a solid foundation for future natural history investigations of CTNNB1 syndrome. Overall, the results highlight craniofacial morphology and selected neuropsychological domains as promising candidates for clinical monitoring, patient stratification and potential therapeutic outcome measures. Moving forward, the systematic integration of genotypic, clinical, developmental and morphometric data will be essential to advance precision phenotyping and to clarify the biological architecture underlying this disorder.

## Statements and declarations

### Data availability statement

The data is not publicly available due to private information from the participants. However, it can be formally requested to the corresponding authors, M.P.S and A.C..

### Funding statement

This study was funded by the Ministry of Sciences, Innovation and Universities of Spain under Grant “Formación Profesorado Universitario” (FPU22/00391 to Mercè Pallarès); “Fundación Inocente Inocente” (Grant FII2024-69); and “Federación Española de Enfermedades Raras” (FEDER) (Grant AI-2023-017). AR-R is funded by Centro de Investigación Biomédica en Red de Enfermedades Raras, CIBERER, Spain. The photographic material was taken under the protocol approved by bioethics committee of Universitat de Barcelona (CBUB) (IRB00003099, CER032514).

### Conflicts of interest disclosure

The authors declare no conflicts of interest.

### Patient and consent statement

All study procedures were in line with the Declaration of Helsinki, complying with all relevant ethical regulations and the protocol was approved by the Ethics Committee of the University of Deusto (ETK-24/23-24) and the Bioethics Committee of the Universitat de Barcelona (IRB03099-CER032514). Written informed consent was obtained from all parents or legal guardians of study participation.

## Acknowledgments

We would like to thank once again the Spanish CTNNB1 Association for always supporting our research. We are also deeply grateful to every family who participated in this study and who fights every day to advance research on CTNNB1 syndrome. All this work is made thanks to you.

## Notes

### Competing Interest Statement

The authors have declared no competing interest.

### Funding Statement

This study was funded by the Ministry of Sciences, Innovation and Universities of Spain under Grant Formacion Profesorado Universitario (FPU22/00391 to Merce Pallares); Fundacion Inocente Inocente (Grant FII2024-69); and Federacion Espanola de Enfermedades Raras (FEDER) (Grant AI-2023-017). AR-R is funded by Centro de Investigacion Biomedica en Red de Enfermedades Raras, CIBERER, Spain.

### Author Declarations

Ethics committee/IRB03099 of the University of Barcelona gave CER032514 approval for this work

## References

Adams, D. C., & Otárola-Castillo, E. (2013). Geomorph: an R package for the collection and analysis of geometric morphometric shape data. Methods in Ecology and Evolution, 4(4), 393–399. 10.1111/2041-210X.12035

Algethami, H., Lim, W. K., Chitayat, D., Tein, I., Fasano, A., & Gorodetsky, C. (2025). Levodopa-Responsive Dystonia Secondary to CTNNB1 Neurodevelopmental Disorder. Movement Disorders Clinical Practice, 12(11), 2012–2014. 10.1002/mdc3.70166

Alvi, A. M., Siuly, S., Wang, H., Wang, K., & Whittaker, F. (2022). A deep learning based framework for diagnosis of mild cognitive impairment. Knowledge-Based Systems, 248, 108815. 10.1016/j.knosys.2022.108815

Ameis, S. H., Ducharme, S., Albaugh, M. D., Hudziak, J. J., Botteron, K. N., Lepage, C., Karama, S., & Brain Development Cooperative Group. (2014). Cortical Thickness, Cortico-Amygdalar Networks, And Externalizing Behaviors In Healthy Children. Biological Psychiatry, 75(1), 65–72. 10.1016/j.biopsych.2013.06.008

Bardua, C., Felice, R. N., Watanabe, A., Fabre, A. C., & Goswami, A. (2019). A Practical Guide to Sliding and Surface Semilandmarks in Morphometric Analyses. Integrative Organismal Biology, 1(1). 10.1093/IOB/OBZ016

Bisgaard, A. M., Schönewolf-Greulich, B., & Gerdes, T. (2021). Decline in gross motor function in adult Rett syndrome. European Journal of Paediatric Neurology, 33, 23–29. 10.1016/j.ejpn.2021.04.004

Bordes Edgar, V., Dorsman, K. A., Horton, D., Messahel, S., & MacDonald, B. (2024). Neuropsychological assessment in rare pediatric neurogenetic disorders: considerations for cross-cultural clinical research. Child Neuropsychology: A Journal on Normal and Abnormal Development in Childhood and Adolescence, 30(6), 900–917. 10.1080/09297049.2023.2283939

Braddock, S. R., Lipinski, R. J., & Carey, J. C. (2020). 40th Annual David W Smith Workshop on Malformations and Morphogenesis: Abstracts of the 2019 Annual Meeting. American Journal of Medical Genetics. Part A, 182(4), 877–942. 10.1002/ajmg.a.61514

Brault, V., Moore, R., Kutsch, S., Ishibashi, M., Rowitch, D. H., McMahon, A. P., Sommer, L., Boussadia, O., & Kemler, R. (2001). Inactivation of the β-catenin gene by Wnt1-Cre-mediated deletion results in dramatic brain malformation and failure of craniofacial development. Development, 128(8), 1253–1264. 10.1242/dev.128.8.1253

Chenn, A. (2008). Wnt/β-catenin signaling in cerebral cortical development. Organogenesis, 4(2), 76–80. 10.4161/org.4.2.5854

Choe, Y., Huynh, T., & Pleasure, S. J. (2014). Neural crest-derived mesenchymal cells require Wnt signaling for their development and drive forebrain morphogenesis. Development, 141(2), 249–260. 10.1242/dev.095612

Cignoni, P., Callieri, M., Corsini, M., Dellepiane, M., Ganovelli, F., & Ranzuglia, G. (2008). MeshLab: an Open-Source Mesh Processing Tool. Sixth Eurographics Italian Chapter Conference, 129–136. 10.2312/LocalChapterEvents/ItalChap/ItalianChapConf2008/129-136

Clayton-Smith, J., Bromley, R., Dean, J., Journel, H., Odent, S., Wood, A., Williams, J., Cuthbert, V., Hackett, L., Aslam, N., Malm, H., James, G., Westbom, L., Day, R., Ladusans, E., Jackson, A., Bruce, I., Walker, R., Sidhu, S., Dyer, C., Ashworth, J., Hindley, D., Arca Diaz, G., Rawson, M., & Turnpenny, P. (2019). Diagnosis and management of fetal valproate spectrum disorder; a consensus statement from the European Reference Network for Congenital Malformations and Intellectual Disability. Orphanet Journal of Rare Diseases, 14, 180. 10.1186/s13023-019-1064-y

de Ligt, J., Willemsen, M. H., van Bon, B. W. M., Kleefstra, T., Yntema, H. G., Kroes, T., Vulto-van Silfhout, A. T., Koolen, D. A., de Vries, P., Gilissen, C., del Rosario, M., Hoischen, A., Scheffer, H., de Vries, B. B. A., Brunner, H. G., Veltman, J. A., & Vissers, L. E. L. M. (2012). Diagnostic Exome Sequencing in Persons with Severe Intellectual Disability. New England Journal of Medicine, 367(20), 1921–1929. 10.1056/NEJMoa1206524

DiLiberti, J. H., Farndon, P. A., Dennis, N. R., & Curry, C. J. (1984). The fetal valproate syndrome. American Journal of Medical Genetics, 19(3), 473–481. 10.1002/ajmg.1320190310

Dong, F., Jiang, J., McSweeney, C., Zou, D., Liu, L., & Mao, Y. (2016). Deletion of CTNNB1 in inhibitory circuitry contributes to autism-associated behavioral deficits. Human Molecular Genetics, 25(13), 2738–2751. 10.1093/hmg/ddw131

Dryden, I., & Mardia, K. (2016). Statistical Shape Analysis, with Applications in R (2nd ed.). 10.1002/9781119072492

Dubruc, E., Putoux, A., Labalme, A., Rougeot, C., Sanlaville, D., & Edery, P. (2014). A new intellectual disability syndrome caused by CTNNB1 haploinsufficiency. American Journal of Medical Genetics Part A, 146(6), 1571–1575. 10.1002/ajmg.a.36484

Dunn, Ll. M., Dunn L.M., & Arribas, D. (2010). PPVT-III Peabody Test de Vocabulario en imágenes. TEA.

Echeverry-Quiceno, L.M., Candelo, E., Gómez, E., Solís, P., Ramírez, D., Ortiz, D., González, A., Sevillano, X., Cuéllar, J.C., Pachajoa, H., & Martínez-Abadías, N. (2023). Population-specific facial traits and diagnosis accuracy of genetic and rare diseases in an admixed Colombian population. Scientific Reports, 13(6869). 10.1038/s41598-023-33374-x

Eklund, H., Findon, J., Cadman, T., Hayward, H., Murphy, D., Asherson, P., Glaser, K., & Xenitidis, K. (2018). Needs of Adolescents and Young Adults with Neurodevelopmental Disorders: Comparisons of Young People and Parent Perspectives. Journal of Autism and Developmental Disorders, 48(1), 83–91. 10.1007/s10803-017-3295-x

Eliasson, A. C., Krumlinde-Sundholm, L., Rösblad, B., Beckung, E., Arner, M., Öhrvall, A. M., & Rosenbaum, P. (2006). The Manual Ability Classification System (MACS) for children with cerebral palsy: scale development and evidence of validity are reliability. Developmental Medicine and Child Neurology, 48(7), 549–554. 10.1111/j.1469-8749.2006.tb01313.x

Elkouby, Y. M., & Frank, D. (2010). Neural crest induction: Wnt/β-catenin signaling. In Wnt/β-Catenin Signaling in Vertebrate Posterior Neural Development (Chapter 8). NCBI Bookshelf. NBK53470.

Ferre-Fernández, M., Murcia-González, M., & Ríos-Díaz, J. (2020). Translation and cross-cultural adaptation of the Gross Motor Function Measure to the Spanish population of children with cerebral palsy. Revista de Neurología, 71(5), 177–185. 10.33588/rn.7105.2020087

Foley, K. R., Downs, J., Bebbington, A., Jacoby, P., Girdler, S., Kaufmann, W. E., & Leonard, H. (2011). Change in Gross Motor Abilities of Girls and Women with Rett Syndrome Over a 3-to 4-Year Period. Journal of Child Neurology, 26(10), 1237–1245. 10.1177/0883073811402688

García-Villamisar, D. (1992). Escala de Clasificación del Autismo Infantil. Acta Pediátrica Española, 50, 383–388.

Gunz, P., & Mitteroecker, P. (2013). Semilandmarks: A Method for Quantifying Curves and Surfaces. Hystrix, the Italian Journal of Mammalogy, 24(1), 103–109. 10.4404/hystrix-24.1-6292

Gunz, P., Mitteroecker, P., & Bookstein, F. L. (2005). Semilandmarks in Three Dimensions. In Modern Morphometrics in Physical Anthropology (pp. 73–98). 10.1007/0-387-27614-9_3

Gurovich, Y., Hanani, Y., Bar, O., Nadav, G., Fleischer, N., Gelbman, D., Basel-Salmon, L., Krawitz, P. M., Kamphausen, S. B., Zenker, M., Bird, L. M., & Gripp, K. W. (2019). Identifying facial phenotypes of genetic disorders using deep learning. Nature Medicine, 25(1), 60–64. 10.1038/s41591-018-0279-0

Hallgrímsson, B., Aponte, J. D., Katz, D. C., Bannister, J. J., Riccardi, S. L., Mahasuwan, N., McInnes, B. L., Ferrara, T. M., Lipman, D. M., Neves, A. B., Spitzmacher, J. A. J., Larson, J. R., Bellus, G. A., Pham, A. M., Aboujaoude, E., Benke, T. A., Chatfield, K. C., Davis, S. M., Elias, E. R., Enzenauer, R. W., French, B. M., Pickler, L. L., Shieh, J. T. C., Slavotinek, A., Harrop, A. R., Tsai, A. C.-H., Wyse, J. P. H., Bernstein, J. A., Sanchez-Lara, P. A., Forkert, N. D., Bernier, F. P., Spritz, R. A., & Klein, O. D. (2020). Automated syndrome diagnosis by three-dimensional facial imaging. Genetics in medicine: official journal of the American College of Medical Genetics, 22(10), 1682–1693. 10.1038/s41436-020-0845-y

Hallgrimsson, B., Percival, C. J., Green, R., Young, N. M., Mio, W., & Marcucio, R. (2015). Morphometrics, 3D Imaging, and Craniofacial Development. Current Topics in Developmental Biology, 115, 561–597. 10.1016/bs.ctdb.2015.09.003

Hamdan, F. F., Srour, M., Capo-Chichi, J. M., Daoud, H., Nassif, C., Patry, L., Massicotte, C., Ambalavanan, A., Spiegelman, D., Diallo, O., Henrion, E., Dionne-Laporte, A., Fougerat, A., Pshezhetsky, A. V., Venkateswaran, S., Rouleau, G. A., & Michaud, J. L. (2014). De Novo Mutations in Moderate or Severe Intellectual Disability. PLoS Genetics, 10(10), e1004772. 10.1371/journal.pgen.1004772

Heredia-Lidón, Á., García-Mascarell, C., Echeverry-Quiceno, L. M., Hostalet, N., Herrera-Escartín, D., González, A., Pomarol-Clotet, E., Fortea, J., Fatjó-Vilas, M., Martínez-Abadías, N. & Sevillano, X. (2025). A Critical Comparison Between Template-Based and Architecture-Reused Deep Learning Methods for Generic 3D Landmarking of Anatomical Structures. In: Wachinger, C., Paniagua, B., Elhabian, S., Luijten, G., Egger, J. (eds) Shape in Medical Imaging. ShapeMI 2024. Lecture Notes in Computer Science, vol 15275. Springer, Cham.

Ho, S. K. L., Tsang, M. H. Y., Lee, M., Cheng, S. S. W., Luk, H. M., Lo, I. F. M., & Chung, B. H. Y. (2022). CTNNB1 Neurodevelopmental Disorder. In GeneReviews®. University of Washington, Seattle.

Hsieh, T. C., Bar-Haim, A., Moosa, S., Ehmke, N., Gripp, K. W., Pantel, J. T., Danyel, M., Mensah, M. A., Horn, D., Rosnev, S., Fleischer, N., Bonini, G., Hustinx, A., Schmid, A., Knaus, A., Javanmardi, B., Klinkhammer, H., Lesmann, H., Sivalingam, S., Kamphans, T., Meiswinkel, W., Ebstein, F., Krüger, E., Küry, S., Bézieau, S., Schmidt, A., Peters, S., Engels, H., Mangold, E., Kreiß, M., Cremer, K., Perne, C., Betz, R. C., Bender, T., Grundmann-Hauser, K., Haack, T. B., Wagner, M., Brunet, T., Bentzen, H. B., Averdunk, L., Coetzer, K. C., Lyon, G. J., Spielmann, M., Schaaf, C. P., Mundlos, S., Nöthen, M. M., & Krawitz, P. M. (2022). GestaltMatcher facilitates rare disease matching using facial phenotype descriptors. Nature Genetics, 54(3), 349–357. 10.1038/s41588-021-01010-x

Hustinx, A., Hellmann, F., Sumer, O., Javanmardi, B., Andre, E., Krawitz, P., & Hsieh, T.-C. (2023). Improving Deep Facial Phenotyping for Ultra-rare Disorder Verification Using Model Ensembles. In Proceedings of the IEEE/CVF Winter Conference on Applications of Computer Vision (pp. 5007–5017). 10.1109/WACV56688.2023.00499

Ji, Y., Hao, H., Reynolds, K., McMahon, M., & Zhou, C. J. (2019). Wnt Signaling in Neural Crest Ontogenesis and Oncogenesis. Cells, 8(10), 1173. 10.3390/cells8101173

Ji, Y., Xia, Q., Zhang, H., Huo, H., Cao, X., Wang, W., & Gu, Q. (2023). Whole Exome Sequencing Identified two Novel Truncation Mutations in the CTNNB1 Gene Associated with Neurodevelopmental Disorder, Language Dysfunction, and Microcephaly in Chinese Children. Child Neurology Open, 10, 2329048X231184184. 10.1177/2329048X231184184

Kaplan E., Goodglass H., & Weintraub S. (2005). Test de Vocabulario de Boston. Panamericana.

Keith, D., & Arellano, I.T. (2012). Sistema de Clasificación de Comunicación Funcional (CFCS) para Personas con Parálisis Cerebral. Instituto Nacional de Rehabilitación

Korkman M., Kirk U., & Kemp S. (2007). NEPSY-II Evaluación Neuropsicológica Infantil. Pearson Educación.

Kuechler, A., Willemsen, M. H., Albrecht, B., Bacino, C. A., Bartholomew, D. W., van Bokhoven, H., van den Boogaard, M. J. H., Bramswig, N., Büttner, C., Cremer, K., Czeschik, J. C., Engels, H., van Gassen, K., Graf, E., van Haelst, M., He, W., Hogue, J. S., Kempers, M., Koolen, D., Monroe, G., de Munnik, S., Pastore, M., Reis, A., Reuter, M. S., Tegay, D. H., Veltman, J., Visser, G., van Hasselt, P., Smeets, E. E. J., Vissers, L., Wieland, T., Wissink, W., Yntema, H., Zink, A. M., Strom, T. M., Lüdecke, H.-J., Kleefstra, T., & Wieczorek, D. (2015). De novo mutations in beta-catenin (CTNNB1) appear to be a frequent cause of intellectual disability: expanding the mutational and clinical spectrum. Human Genetics, 134 (1), 97–109. 10.1007/s00439-014-1498-1

Kuwahara, Y., Yonezawa, M., Miya, H., Moroto, M., & Iehara, T. (2025). Behavioral problems in adolescents and young adults with neurodevelopmental disorders. Pediatrics International: Official Journal of the Japan Pediatric Society, 67(1), e70115. 10.1111/ped.70115

Kwiatkowski, A. V., & Weis, W. I., & Nelson, W. J. (2007). Catenins: Playing both sides of the synapse. Neuron, 19(5), 551–556. 10.1016/j.ceb.2007.08.005

Lee, E., Choi, J. Y., & Yang, S. S. (2025). Case report: Discovery of novel *CTNNB1* mutations and comparison of clinical characteristics in two patients with NEDSDV. Frontiers in Genetics, 16, 1502756. 10.3389/fgene.2025.1502756

Lee, J., Yoo, J., Lee, S., & Jang, D. H. (2023). CTNNB1-related neurodevelopmental disorder mimics cerebral palsy: case report. Frontiers in Pediatrics, 11, 1201080. 10.3389/fped.2023.1201080

Levy, D., Ronemus, M., Yamrom, B., Lee, Y. H., Leotta, A., Kendall, J., Marks, S., Lakshmi, B., Pai, D., Ye, K., Buja, A., Krieger, A., Yoon, S., Troge, J., Rodgers, L., Iossifov, I., & Wigler, M. (2011). Rare De Novo and Transmitted Copy-Number Variation in Autistic Spectrum Disorders. Neuron, 70(5), 886–897. 10.1016/j.neuron.2011.05.015

Lindsay, P., Virella, D., Mjøen, T., da Graça-Andrada, M., Murray, J., Colver, A., Himmelmann, K., Rackauskaite, G., Greitane, A., Prasauskiene, A., Andersen, G., & de la Cruz, J. (2013). Development of The Viking Speech Scale to classify the speech of children with cerebral palsy. Research in Developmental Disabilities, 34(10), 3202–3210. 10.1016/j.ridd.2013.06.035

Maguschak, K. A., & Ressler, K. J. (2012). The dynamic role of β-catenin in synaptic plasticity. Neuropharmacology, 62(1), 78–88. 10.1016/j.neuropharm.2011.08.032

Marchini, M., Hu, D., Lo Vercio, L., Young, N. M., Forkert, N. D., Hallgrímsson, B., & Marcucio, R. (2021). Wnt Signaling Drives Correlated Changes in Facial Morphology and Brain Shape. Frontiers in Cell and Developmental Biology, 9, 644099. 10.3389/fcell.2021.644099

Marcucio, R., Hallgrimsson, B., & Young, N. M. (2015). Facial Morphogenesis: Physical and Molecular Interactions Between the Brain and the Face. Current topics in developmental biology, 115, 299–320. 10.1016/bs.ctdb.2015.09.001

Martínez-Abadías, N., Heuzé, Y., Wang, Y., Jabs, E. W., Aldridge, K., & Richtsmeier, J. T. (2011). FGF/FGFR Signaling Coordinates Skull Development by Modulating Magnitude of Morphological Integration: Evidence from Apert Syndrome Mouse Models. PloS ONE, 6(10), e26425. 10.1371/journal.pone.0026425

Miroševič, Š., Khandelwal, S., Amerson, E., Parks, E., Parks, M., Cochran, L., González Hernández, A., Ferraro, M., Lisowski, L., Perez-Iturralde, A., Chung, W., Jacob, M. H., Žakelj, N., Lainšček, D., Forstnerič, V., Sušjan, P., Maruna, M., Jerala, R., & Osredkar, D. (2025). Paving the way toward treatment solutions for CTNNB1 syndrome: a patient organization perspective. Therapeutic Advances in Rare Disease, 6, 26330040251318355. 10.1177/26330040251318355

Miroševič, Š., Khandelwal, S., Sušjan, P., Žakelj, N., Gosar, D., Forstnerič, V., Lainšček, D., Jerala, R., & Osredkar, D. (2022). Correlation between Phenotype and Genotype in CTNNB1 Syndrome: A Systematic Review of the Literature. International Journal of Molecular Sciences, 23(20), 12564. 10.3390/ijms232012564

Nunes-Xavier, C. E., Pallarès-Sastre, M., Rodríguez-Ramos, A., Bañuelos, S., Cortajarena, I., Cavaliere, F., Ruiz-Espinoza, C., Llano-Rivas, I., García, M., Amayra, I., & Pulido, R. (2025). Genotype-phenotype characterization and functional reconstitution of pathogenic β-catenin variants from CTNNB1 syndrome patients. PLoS Genetics, 21(10), e1011907. 10.1371/journal.pgen.1011907

Pagerols, M., Bosch, R., Prat, R., Pagespetit, È., Cilveti, R., Chaparro, N., Esteve. A., & Casas, M. (2023). The Sleep Disturbances for Children: psychometric properties and prevalence of sleep disorders in Spanish children aged 6-16 years. Journal of Sleep Research, 32(4), e13871. 10.1111/jsr.13871

Pallarès-Sastre, M., Amayra, I., Pulido, R., Nunes-Xavier, C. E., Bañuelos, S., Cavaliere, F., & García, M. (2025a). Cognitive and Adaptive Functioning of CTNNB1 Syndrome Patients: A Comparison With Autism Spectrum Disorder and Cerebral Palsy. Journal of Intellectual Disability Research, 69, 558–568. 10.1111/jir.13235

Pallarès-Sastre, M., Amayra, I., Pulido, R., Nunes-Xavier, C. E., Bañuelos, S., Cavaliere, F., & García, M. (2025b). Novel CTNNB1 Gene Variants in Spanish CTNNB1 Syndrome Patients: Clinical and Psychological Manifestations. Journal of Autism and Developmental Disorders. 10.1007/s10803-025-06829-5

Pallarès-Sastre, M., Amayra, I., Salgueiro, M., Villanueva-Viar, E., Lasa-Aranzasti, A., & García, M. (2025). A Systematic Review of Cognitive and Behavioural Symptoms in CTNNB1 Syndrome. Neuropsychology Review. 10.1007/s11065-025-09660-y

Paulsen, R. R., Juhl, K. A., Haspang, T. M., Hansen, T., Ganz, M., & Einarsson, G. (2019). Multiview Consensus CNN for 3D Facial Landmark Placement. In Jawahar, C., Li, H., Mori, G., Schindler, K. (eds) Computer Vision – ACCV 2018. ACCV 2018. Lecture Notes in Computer Science, 11361, 706–719. Springer, Cham. 10.1007/978-3-030-20887-5_44

Pereña, J., & Santamaría, P. (2005). Cuestionario de Comunicación Social (SCQ). TEA Ediciones.

Pipo-Deveza, J., Fehlings, D., Chitayat, D., Yoon, G., Sroka, H., & Tein, I. (2018). Rationale for dopa-responsive CTNNB1/ β-catenin deficient dystonia. Movement Disorders, 33(4), 656–657. 10.1002/mds.27320

Schatz, J., & Hamdan-Allen, G. (1995). Effects of age and IQ on adaptive behavior domains for children with autism. Journal of Autism and Developmental Disorders, 25(1), 51–60. 10.1007/BF02178167

Schlager, S., & Rüdell, A. (2017). Sexual Dimorphism and Population Affinity in the Human Zygomatic Structure—Comparing Surface to Outline Data. Anatomical Record, 300(1), 226–237. 10.1002/ar.23450

Schlager, S., Jefferis, G., & Ian, D. (2018). Package “Morpho” Type Package Title Calculations and Visualisations Related to Geometric Morphometrics.

Sellers, D., Mandy, A., Pennington, L., Hankins, M., & Morris, C. (2013). Development and reliability of a system to classify eating and drinking ability of people with cerebral palsy. Developmental Medicine Child Neurology, 15(3), 245–251. 10.1111/dmcn.12352

Sparrow S. S., Cicchetti D. V., & Balla D. A. (2016). Vineland 3. Spanish parent/caregiver form Comprehensive version. Pearson.

Sudnawa, K. K., Garber, A., Cohen, R., Calamia, S., Kanner, C. H., Montes, J., Bain, J. M., Fee, R. J., & Chung, W. K. (2024). Clinical phenotypic spectrum of CTNNB1 neurodevelopmental disorder. Clinical Genetics, 105(5), 523–532. 10.1111/cge.14487

Sutton, G., Kelsh, R. N., & Scholpp, S. (2021). The Role of Wnt/β-Catenin Signaling in Neural Crest Development in Zebrafish. Frontiers in Cell and Developmental Biology, 9, 782445. 10.3389/fcell.2021.782445

Tang, S. J. (2014). Synaptic activity-regulated Wnt signaling in synaptic plasticity, glial function and chronic pain. CNS & Neurological Disorders Drug Targets, 13(5), 737–744. 10.2174/1871527312666131223114457

Tucci, V., Kleefstra, T., Hardy, A., Heise, I., Maggi, S., Willemsen, M. H., Hilton, H., Esapa, C., Simon, M., Buenavista, M.-T., McGuffin, L. J., Vizor, L., Dodero, L., Tsaftaris, S., Romero, R., Nillesen, W. N., Vissers, L. E. L. M., Kempers, M. J., Vulto-van Silfhout, A. T., Iqbal, Z., Orlando, M., Maccione, A., Lassi, G., Farisello, P., Contestabile, A., Tinarelli, F., Nieus, T., Raimondi, A., Greco, B., Cantatore, D., Gasparini, L., Berdondini, L., Bifone, A., Gozzi, A., Wells, S., & Nolan, P. M. (2014). Dominant β-catenin mutations cause intellectual disability with recognizable syndromic features. The Journal of Clinical Investigation, 124(4), 1468–1482. 10.1172/JCI70372

Tucci, V., Kleefstra, T., Hardy, A., Heise, I., Maggi, S., Willemsen, M. H., Hilton, H., Esapa, C., Simon, M., Buenavista, M. T., McGuffin, L. J., Vizor, L., Dodero, L., Tsaftaris, S., Romero, R., Nillesen, W. N., Vissers, L. E. L. M., Kempers, M. J., Vulto-van Silfhout, A. T., Iqbal, Z., Orlando, M., Maccione, A., Lassi, G., Farisello, P., Contestabile, A., Tinarelli, F., Nieus, T., Raimondi, A., Greco, B., Cantatore, D., Gasparini, L., Berdondini, L., Bifone, A., Gozzi, A., Wells, S., & Nolan, P. M. (2014). Dominant β-catenin mutations cause intellectual disability with recognizable syndromic features. The Journal of Clinical Investigation, 124(4), 1468–1482. 10.1172/JCI70372

Verhoeven, W. M. A., Egger, J. I. M., Jongbloed, R. E., Meijer van Putten M., de Bruin-van Zandwijk, M., Zwemer, A.-S., Pfundt, R., & Willemsen, M. H. (2020). *A de novo CTNNB1 novel splice variant in an adult female with severe intellectual disability* (case report). International Medical Case Reports Journal, 13, 487–492. 10.2147/IMCRJ.S270487

Verhoeven, W. M. A., Egger, J. I. M., Jongbloed, R. E., van Putten, M. M., de Bruin-van Zandwijk, M., Zwemer, A. S., Pfundt, R., & Willemsen, M. H. (2020). A de novo CTNNB1 Novel Splice Variant in an Adult Female with Severe Intellectual Disability. International Medical Case Reports Journal, 13, 487–492. 10.2147/IMCRJ.S270487

Wadden, J. J. (2022). Defining the undefinable: The black box problem in healthcare artificial intelligence. Journal of Medical Ethics, 48(10), 764–768. 10.1136/medethics-2021-107529

Walpert, M., Zaman, S., & Holland, A. (2021). A Systematic Review of Unexplained Early Regression in Adolescents and Adults with Down Syndrome. Brain Sciences, 11(9), 1197. 10.3390/brainsci11091197

Wechsler D. & Naglieri J.A. (2011). Escala No Verbal de Aptitud Intelectual de Wechsler. Medida no verbal de la inteligencia general. Pearson.

Winczewska-Wiktor, A., Badura-Stronka, M., Monies-Nowicka, A., Nowicki, M. M., Steinborn, B., Latos-Bieleńska, A., & Monies, D. (2016). A de novo CTNNB1 nonsense mutation associated with syndromic atypical hyperekplexia, microcephaly and intellectual disability: a case report. BMC Neurology, 16, 35. 10.1186/s12883-016-0554-y

Winter, R. M., Donnai, D., Burn, J., & Tucker, S. M. (1987). Fetal valproate syndrome: is there a recognisable phenotype?. Journal of medical genetics, 24(11), 692–695. 10.1136/jmg.24.11.692

Yan, D., Sun, Y., Xu, N., Yu, Y., Zhan, Y., & Mainland Chinese League of NEDSDV Rare Disease (2022). Genetic and clinical characteristics of 24 mainland Chinese patients with CTNNB1 loss-of-function variants. Molecular Genetics & Genomic Medicine, 10(11), e2067. 10.1002/mgg3.2067

Youngs, E. L., Hellings, J. A., & Butler, M. G. (2011). A clinical report and further delineation of the 14q32 deletion syndrome. Clinical Dysmorphology, 20(3), 143–147. 10.1097/MCD.0b013e3283438200

Žakelj, N., Gosar, D., Miroševič, Š., Sanders, S. J., Ljungdahl, A., Kohani, S., Huang, S., Leong, L. I., An, Y., Teo, M. J., Moultrie, F., Jerala, R., Lainšček, D., Forstnerič, V., Sušjan, P., Lisowski, L., Perez-Iturralde, A., Mrak, J. O., Chan, H. Y. E., & Osredkar, D. (2025). Genotypic, functional, and phenotypic characterization in CTNNB1 neurodevelopmental syndrome. HGG Advances, 6(4), 100483. 10.1016/j.xhgg.2025.100483

